# Stakeholder perspectives on community-based nutrition interventions for child stunting in Rwanda: A qualitative comparative study of caregivers, health workers and health service managers

**DOI:** 10.64898/2026.11.16.26368249

**Authors:** Seleman Ntawuyirushintege, Nouh Saad Mohamed, Ayman Ahmed, Stephen Rulisa, Eric Remera, Kaspar Wyss

**Affiliations:** College of Medicine and Health Sciences, University of Rwanda, Kigali, Rwanda; Swiss Centre for International Health, Swiss Tropical and Public Health Institute (Swiss TPH), Allschwil, Switzerland; Medical Faculty, University of Basel, Basel, Switzerland; Pan-Africa One Health Institute (PAOHI), Kigali, Rwanda; Institute of Endemic Diseases, University of Khartoum, Khartoum, Sudan; Division of Research Innovation and Data Science, Rwanda Biomedical Centre, Kigali, Rwanda

**Author notes:** Corresponding author (SN).

## Abstract

Stunting among children under 2 years old remains a major public health concern in Rwanda despite the efforts to reduce its prevalence. Community-based nutrition interventions, including food supplementation, growth monitoring, screening and referral, are widely delivered, yet high coverage has not consistently translated into lower stunting prevalence. This study explores caregivers, community health workers (CHWs), and health service managers experiences’ and understanding of these interventions.

A qualitative descriptive study was conducted in the areas of Bugeshi (Rubavu district) and Rugarama (Nyarugenge district) health centres, purposively selected for their contrasting district stunting trajectories. We conducted six focus group discussions with caregivers and CHWs (48 participants) and 21 in-depth interviews with facility-based nutritionists and environmental health officers, local health service managers, and national, international and private-sector programme managers and experts. Data analysis using reflexive thematic analysis showed seven emerging themes, including competing explanations for persistent stunting; experiences of receiving and using food supplements; the emotional and social dimensions of malnutrition; the central but under-resourced, and at times unsafe, role of CHWs; difficulties in reaching eligible children; governance and financial sustainability; and supply chain constraints. All groups valued food supplementation and shared an understanding of stunting. Caregivers and frontline CHWs more often described problems at the point of delivery such as eligible children being missed, supplements being shared with siblings and other household members or being unavailable, and CHWs facing hostility during periods of stock-outs. Managers and programme experts emphasised financing, procurement and distribution as essential pillar for intervention success.

Community-based nutrition interventions were widely valued, but participants described a substantial gap between programme design and day-to-day implementation. Strengthening implementation will require attention to the working conditions of CHWs, household economic realities, programme eligibility criteria, supply and financing systems, and routine use of caregiver and CHWs feedback in programme monitoring.

## Introduction

Childhood stunting is a chronic form of undernutrition that typically originates in the first 1,000 days from conception to a child’s second birthday. Stunting is associated with impaired cognitive development, reduced adult productivity and greater susceptibility to disease, and is widely regarded as a marker of a population’s human capital [1].

Rwanda has substantially reduced childhood stunting through sustained investment under successive national development strategies [2–4]. The Second National Strategy for Transformation (NST2, 2024–2029) sets a national target of 15% by 2029 [5].

In Rwanda, among children under 5 years stunting prevalence fell from 51% in 2005 to 33% in 2019–20 [6] and to 27% in 2025 [7]. These gains nevertheless remain uneven. In the 2025 survey, prevalence ranged from less than 10% in Nyarugenge district in Kigali to more than 30% in 11 districts, most of them located in the Western Province [7]. Routine screening of children aged 2 years between 202-2024 showed a similar pattern with a national prevalence in this age group falling from 33% to 22%, but district trends diverged markedly, and the smallest relative reductions were recorded in districts of the Western Province [8], highlighting the need for understanding community and stakeholder perspectives to explain these differences.

Rwanda’s response although embedded in national nutrition and health sector policies it combines child-focused nutrition services with community-based delivery [9–11]. Micronutrient powders have been distributed to children aged 6–23 months under the “Ongera” programme since 2012 [12]. Since 2017, locally produced fortified blended foods, such as “Shisha Kibondo” and “SOSOMA” (a sorghum–soy–maize flour), have been distributed monthly through health centres, alongside advocacy and counselling on their preparation and on infant and young child feeding [12,13]. The eligibility for fortified blended food and other social protection services has historically been combined the household’s socio-economic category under the national Ubudehe classification, a community-based poverty-ranking system introduction. Ubudehe originally comprised six categories, which were consolidated into four numbered categories in 2015 and revised in 2020 into five lettered categories (A–E), ranging from the highest-income households (A) to the most vulnerable (E) [14,15]. Owing to inclusion and exclusion errors in beneficiary selection, the government discontinued the use of Ubudehe categories for targeting in 2023. In February 2024, it was replaced by the Imibereho Social Registry, which classifies households primarily on the basis of income, supplemented by asset data such as land and vehicle ownership, drawn from linked administrative databases [14,16]. Individual programmes apply their own thresholds to registry data. For community-based health insurance, for example, households are assigned to one of five income-based contribution categories, ranging from Category I (no income, fully subsidised by the government) to Category V (monthly income above 120,000 Rwandan francs) [17–19].

Implementation of stunting reduction interventions relay mostly on Community Health Workers (CHWs) who carry out routine growth monitoring, home visits and household counselling, run cooking demonstrations in village nutrition kitchens, and refer children with growth faltering or signs of malnutrition to the health centre. At the health centre, nutritionists or nurses assess the child and, where eligibility criteria are met, enrol the child for food supplementation or for treatment of acute malnutrition. Data from these services are recorded in health centre registers and in an electronic individual tracking system (e-Tracker) [20].

Nutritional status is also assessed in mass screening of children by CHWs during the twice-yearly national Maternal and Child Health Weeks and coordinated from village to district level [8,9,11]. Several donor-funded projects supported the implementation of stunting prevention interventions such as the USAID-funded Gikuriro programme (2015–2020) which combined village nutrition schools with savings groups and water, sanitation and hygiene promotion, and trained volunteer “lead mothers” to lead peer learning and cooking demonstrations for other mothers in their village [21]. Also, the World Bank-supported Stunting Prevention and Reduction Project, approved in 2018, provided additional financing for nutrition services [22,23]. Although the coverage of these services was high, in a national survey in 2023, despite 94% of children aged 6–23 months received food supplements, yet 1 in 5 children aged 2 years were still stunted in 2024 [8]. The coexistence of near-universal supplementation coverage and persistently high stunting prevalence suggests that coverage alone does not reflect the quality, continuity or contextual appropriateness of interventions, and highlights the need to understand how programmes are implemented and experienced at local level. Implementation depends not only on the availability of services but also on how they are delivered, accessed, understood and accepted. However, little is known, in Rwanda or in comparable programmes in the region about how these interventions are experienced and enacted by the different actors involved [24–26]. Therefore, this study is aimed at understanding the perspectives of stakeholders, workers delivering nutrition services, in particularly CHWs, facility-based nutritionists, and health service managers, together with international and private-sector programme managers to explore the barriers of interventions’ effectiveness and their implications in programme implementation.

## Materials and methods

### Design

This qualitative descriptive study combined focus group discussions (FGDs) and in-depth interviews (IDIs), analysed using inductive reflexive thematic analysis [27] supported by elements of the Framework Method [28]. The study is reported in accordance with the Consolidated Criteria for Reporting Qualitative Research (COREQ) [29] (S1 Checklist).

### Study setting and site selection

Site selection was informed by a previous analysis of routine national screening data for children aged 2 years [8]. In the period 2020 to 2024, stunting prevalence in Rubavu district, which includes Bugeshi sector, fell from 41% to 35%. This decrease of 6 percentage points corresponds to a relative reduction of 14.6%, one of the 2 smallest relative reductions among Rwanda’s 30 districts. In contrast, prevalence in Nyarugenge district, which includes Rugarama sector, fell from 23% to 6%, a decrease of 17 percentage points and a relative reduction of 73.9%, one of the largest reductions nation-wide [8]. Therefore, those 2 settings were purposively selected as they provide contrasting trends rather than on baseline prevalence alone. Bugeshi is predominantly rural, agrarian and water-scarce area bordering the Democratic Republic of the Congo (DRC), while Rugarama is a mixed urban and peri-urban setting.

### Participants and sampling

Participants were purposively sampled from 3 stakeholder groups. Group 1 comprised caregivers of children under 2 years who were using or had used community-based nutrition services. Group 2 comprised health workers delivering nutrition services, including CHWs, facility and district hospital nutritionists, and Community Environmental Health Officers (CEHOs). Group 3 comprised health service managers and programme experts involved in overseeing, coordinating or supporting nutrition interventions at local and national levels. Those at local level included health centre managers, Early Childhood Development (ECD) coordinators and sector and district officials. Those at national level included representatives of government agencies, United Nations agencies, an international nutrition organisation and a private-sector food supplement manufacturer, and 2 independents public health consultants.

Caregivers and CHWs perspectives were explored through FGDs, which suited the exploration of shared community experiences. At each site, 2 sex-specific caregiver FGDs (1 female, 1 male) were held. In addition, 1 FGD was organised with CHWs at each site and included both female and male CHWs. Data collection for other health workers and for local and national managers and experts was done through IDIs, with a total of 21 interviews conducted. IDI participants were sampled across facility, sector, district and national levels until additional interviews yielded no substantially new issues [30]. Caregivers and CHWs were recruited through health centre registers, and other participants through institutional contacts. All those approached participants agreed to participate and provided informed consent.

### Data collection

Data were collected between May and July 2026 by trained interviewers using semi-structured topic guides (S1 Text) covering perceptions of stunting and its causes, experiences of food supplementation and nutrition services, community engagement, programme monitoring and sustainability, and equity and access. Guides were adapted to each respondent group around a common core of questions to allow comparison across stakeholder groups.

FGDs were conducted in Kinyarwanda in a private room at the health centre and lasted approximately 1 hour. Health centre staff did not take part in caregiver FGDs. Each FGD was facilitated by a moderator and a note-taker. IDIs with facility, sector and district staff took place in the 2 study districts, and IDIs with national, international and private-sector participants took place in Kigali. Sessions were audio-recorded with participants’ consent.

### Data analysis

Recordings were transcribed verbatim, translated into English, quality-checked and analysed in ATLAS.ti version 24. The analysis combined the stages of the Framework Method (familiarisation, coding, framework development, indexing, charting and interpretation) with Braun and Clarke’s iterative, reflexive approach to theme development [27,28]. Codes and themes were developed inductively, without a pre-existing codebook or theoretical framework.

To compare stakeholder groups, transcripts were charted by respondent category in the framework matrix, allowing each theme to be examined for convergence and divergence across caregiver, health worker and manager accounts (code frequencies by group are given in S1 Table). Differences between female and male caregiver FGDs, and inconsistencies within the same professional cadre, were treated as findings rather than noise.

For analysing equity of access to food supplementation and related nutrition services, we focused on horizontal equity, whereby children with equal need should have equal access to services [31,32].

### Ethics statement

The study was approved by the Institutional Review Board of the College of Medicine and Health Sciences, University of Rwanda (Approval Notice No. 452/CMHS IRB/2026; approved 27 April 2026). All participants gave written informed consent, and transcripts were de-identified before analysis.

### Inclusivity in global research

Additional information regarding the ethical, cultural and scientific considerations specific to inclusivity in global research is included in the Supporting Information (S2 Checklist).

## Results

### Participant characteristics and overview of themes

In total, 27 sessions were analysed: 6 FGDs (48 participants) and 21 IDIs, representing 69 participants (Table 1). Caregivers were predominantly married, lived in households of 3 to 7 members and mostly worked in informal or agrarian occupations. CHWs reported between 3 months to 28 years of experience. Managers and programme experts held qualifications ranging from secondary school to Doctor of Philosophy degree.

**Table 1.** Characteristics of study participants by stakeholder group.

| Stakeholder group | Method (n sessions) | Participants (n) | Female / male (n) | Age in years, mean (range) | Other characteristics |
|---|---|---|---|---|---|
| Caregivers, Bugeshi | 2 FGDs: 1 female, 1 male | 17 | 9 / 8 | 33.4 (25–45) | All married; household size 3–7; mainly agrarian or informal occupations |
| Caregivers, Rugarama | 2 FGDs: 1 female, 1 male | 15 | 8 / 7 | 38.7 (25–63) | Predominantly married (14/15); household size 3–6; mainly informal occupations |
| Community Health Workers, Bugeshi | 1 FGD | 8 | 4 / 4 | 45.1 (28–66) | Experience as CHW: 3 months to 20 years |
| Community Health Workers, Rugarama | 1 FGD | 8 | 7 / 1 | 49.5 (36–65) | Experience as CHW: 4 to 28 years |
| Other health workers (facility and hospital nutritionists, CEHOs) | IDI (5) | 5 | 2 / 3 | 40.2 (26–52) | Highest qualification: A1 advanced diploma to bachelor's degree; experience 2 to 30 years |
| Local health service managers (health centre, ECD, sector and district level) | IDI (8) | 8 | 1 / 7 | 41.4 (33–55) | Highest qualification: secondary (Senior 6) to master's degree; experience 1 to 22 years |
| National, international and private-sector programme managers and experts | IDI (8) | 8 | 4 / 4 | 41.8 (38–51) | Government agencies (2), UN agencies (2), international nutrition organisation (1), independent researchers (2), private sector (1); qualifications master's to PhD; experience 8 to 20 years |
| **Total** | **6 FGDs, 21 IDIs** | **69** | **35 / 34** | **40.1 (25–66)** |  |

CEHO, Community Environmental Health Officer; CHW, Community Health Worker; ECD, Early Childhood Development; FGD, focus group discussion; IDI, in-depth interview; UN, United Nations. Other health workers and local managers together account for 13 IDIs. To protect anonymity, organisations are grouped by type and individual job titles are not reported.

Seven themes emerged as central aspects from the FGDs and IDIs: (1) competing explanations of stunting; (2) experiences of receiving and using food supplements; (3) the social dimensions of malnutrition; (4) the central but under-resourced role of CHWs; (5) difficulties in reaching eligible children; (6) governance, monitoring and financial sustainability; and (7) supply chain constraints and stock-outs (Table 2).

**Table 2.** Key issues emerging in each stakeholder group, and points of convergence, by theme.

| **Theme (shared view across groups)** | **Caregivers** | **Health workers (including CHWs)** | **Health service managers and programme experts** |
|---|---|---|---|
| 1. Explanations of stunting<br>*Shared: biomedical first-1,000-days framework* | Underlying illness (e.g., tuberculosis, cleft palate) initially taken for malnutrition; household finances | Caregiver knowledge and behaviour (especially Bugeshi); limited caregiving time; alcohol and domestic conflict | Land ownership concentrated in fewer hands, flooding and affordability rather than caregiver knowledge |
| 2. Food supplements<br>*Shared: supplementation valued; sharing within the household linked to food insecurity* | Children's recovery; relapse after food support ended; opportunity costs of attending services (female FGDs) | Supplements shared with other household members or occasionally sold; early cessation of breastfeeding and avoidance of family planning to keep eligibility; abrupt end of support after household reclassification; rigid distribution schedules | Demand from families whose children do not meet eligibility criteria |
| 3. Social dimensions<br>*Shared: diagnosis can bring shame* | Gossip deters attendance; mutual support between households; resentment among non-beneficiaries (male FGD, Bugeshi) | CHWs delivering supplements privately; wealthier households denying a child's malnutrition or using services secretly | Wealthier households denying a child's malnutrition or avoiding services |
| 4. Community Health Workers<br>*Shared: CHWs most trusted and central actors* | CHWs declining to refer children to the health centre; alleged under-reporting of malnutrition by CHWs | Heavy workload; shortages of anthropometric equipment (height boards, MUAC tapes, weighing scales); unpaid work; insults and threats during stock-outs | Corroboration of tensions with caregivers during stock-outs |
| 5. Reaching eligible children<br>*Shared: formal eligibility criteria exist* | Female FGDs: perceived favouritism in referral and enrolment; poor urban households missed. Male FGD, Rugarama: system seen as fair | Rationing by distance during shortages; poor households missed in wealthier urban neighbourhoods; unmet need among marginalised communities | Confidence that all eligible children are reached; exclusion of eligible children at the point of delivery not mentioned |
| 6. Governance, monitoring and financing<br>*Shared: concern about donor dependence* | Alleged under-reporting of poverty and malnutrition by local leaders and CHWs | Loss of the lead mother model after donor funding ended; doubts about the accuracy of e-Tracker data | Unpredictable funding cycles; more confidence than frontline respondents in the accuracy of routine data |
| 7. Supply chain and stock-outs<br>*Shared: supply reliability a main constraint* | Recurrent stock-outs | Recurrent stock-outs of weeks to months; consequences for families and CHWs | Procurement delays, national bottlenecks and last-mile constraints; coverage dips tracking stock-outs; production concentrated in Kigali seen as a source of supply vulnerability |

CHW, Community Health Worker; FGD, focus group discussion; MUAC, mid-upper arm circumference.

#### Competing explanations of stunting

All 3 stakeholder groups described stunting through the same biomedical framework centred on the first 1,000 days of life, expressed in similar terms by senior managers, by CHWs, including those only a few months in post, and by caregivers. Participants differed, however, in explaining why stunting persists. Health workers and local managers in Bugeshi often attributed it to caregivers’ “poor understanding”, particularly in an area they regarded as agriculturally productive. Others offered structural explanations. A district-level manager described land ownership becoming increasingly concentrated in fewer hands, which left some households farming land owned by others, and identified flooding as a contributor to food insecurity. National participants likewise emphasised household economic constraints; a public health economist considered affordability rather than palatability is the main reason for low uptake of food supplements.

A further explanation concerned caregiving time. In Bugeshi, participants described mothers spending long hours in small-scale cross-border trade with the DRC; in Rugarama, they described children left in the care of hired domestic workers. At both sites, health workers and managers observed that income did not necessarily translate into adequate time or care for young children. Household alcohol use and domestic conflict were also described as pathways to child undernutrition. Water scarcity and cultural feeding taboos were also mentioned, and helminth infections were raised as a concern in Bugeshi.

Caregivers in Rugarama further described children presumed to be malnourished if were found to have an underlying condition, such as tuberculosis or cleft palate, or whose mother had breast cancer. These accounts challenge a purely behavioural reading of growth faltering and show the value of clinical assessment for children identified through growth monitoring.

#### Experiences of receiving and using food supplements

Food supplementation was valued in all stakeholder groups and described as beneficial, and in some cases clinically effective, when used as intended. Caregivers described their children’s recovery, and health workers and managers described demand for supplements from families whose children did not meet the eligibility criteria. These benefits were repeatedly interrupted by stock-outs, and rigid distribution schedules could delay access.

Participants in all groups described families sharing supplements intended for the eligible child with siblings and other household members, and occasionally selling them, and mostly linked this to household food insecurity rather than to a lack of understanding.

Some health workers reported that caregivers had stopped breastfeeding early, or avoided family planning, to keep a child eligible for supplementation. Food support ends when the child reaches the upper age limit or when the household is reclassified into a socio-economic category that is no longer eligible. Health workers reported that the end of support at the age limit was increasingly preceded by preparation of the caregiver, whereas the end of support following reclassification of the household could be abrupt. Caregivers, particularly women, described children relapsing after food support ended when household difficulties had not been resolved. Female caregivers also described the opportunity costs of attending services, such as lost working time.

#### The social dimensions of malnutrition: stigma, shame and solidarity

Caregivers described a diagnosis of stunting or malnutrition as emotionally significant, often involving embarrassment and fear of judgement. Health workers at both sites described CHWs collecting supplements for families who did not want to be seen receiving them and delivering them privately.

Health workers and managers independently described some wealthier households denying that their child was malnourished or avoiding services, including caregivers who rejected a diagnosis or used services “secretly”. Caregivers identified gossip as a deterrent. Several participants who described these behaviours also stated that “there is no stigma” in their community. This suggests that stigma was understood narrowly as open discrimination, and that shame, concealment and avoidance were not recognised as forms of stigma.

Caregivers also described mutual support, including neighbours donating milk, savings-based cooperative schemes and families whose children had recovered serving as examples. In contrast, male caregivers in Bugeshi described resentment among some households that did not receive supplements, including reports of parents giving their own children less food. Although not widespread, these accounts point to a little-examined risk of means-tested nutrition programmes.

#### Community Health Workers: central actors under pressure

All 3 stakeholder groups described CHWs as the most trusted and central actors in the nutrition system, a centrality that came with a heavy workload. CHWs reported chronic shortages of anthropometric equipment, such as height boards, mid-upper arm circumference tapes and weighing scales, and continued reliance on unpaid volunteer labour despite Rwanda’s performance-based financing scheme for CHWs.

CHWs in Bugeshi described threats to their personal safety. CHWs identify children at risk and mobilise caregivers to attend the health centre or village nutrition kitchen; when caregivers found no commodities available, their frustration was directed at the CHWs, the most visible local representative of the programme. CHWs described being insulted, threatened and refused entry to homes. Supply disruptions thus affected not only service availability but also the safety and working relationships of frontline workers.

Caregivers in Rugarama described CHWs deciding not to refer to the health centre children whom caregivers believed needed assessment, effectively acting as gatekeepers to services. Caregivers also alleged that some CHWs and local leaders under-reported cases of malnutrition to protect their reputation. Frontline discretion thus shaped who was identified, referred and reached.

#### Difficulties in reaching eligible children

Equity concerns related mainly to horizontal equity, that is, whether all children meeting the eligibility criteria were identified and served. Managers and programme experts generally expressed confidence that services reached all eligible children. This confidence was less evident among health workers and female caregivers, who were closer to day-to-day delivery. Female caregivers perceived favouritism in which children were referred and enrolled and described poor households in urban neighbourhoods being missed. In Bugeshi, when stock was insufficient for all enrolled children, households closer to the health centre were more likely to receive supplements than those farther away. Stock-outs thus translated directly into unequal access among children with the same need.

The clearest equity concern emerged in Rugarama, where health workers and caregivers independently described poor households being overlooked in wealthier urban neighbourhoods. Outreach concentrated in visibly poor villages could miss poor households in or near wealthier neighbourhoods, and gated compounds and households employing domestic workers were described as barriers to identifying eligible children.

In Bugeshi, managers and health workers described substantial unmet need, and visible frustration, among historically marginalised communities. Inequity can therefore arise not only from formal eligibility criteria but also from how vulnerability is recognised and translated into outreach.

National and international participants described targeting based on formal vulnerability criteria, but none mentioned the exclusion of eligible children at the point of delivery described by health workers and caregivers, suggesting a gap between targeting design and its visibility in practice at national level.

At both sites, female caregiver FGDs gave more detailed and critical accounts of programme barriers, whereas male FGDs more often framed household decisions around the father’s role as provider and judged the distribution of services as fairer.

#### Governance, monitoring and financial sustainability

Participants in all groups were concerned about dependence on external funding. They attributed the end of 2 valued donor-supported initiatives to reduced funding. National and international participants described unpredictable, short or poorly aligned funding cycles as a constraint on multi-year planning. Rather than describing domestic resources as already compensating for the loss of donor funding, they presented greater domestic financing and local production as needs for the future.

#### Supply chain constraints and stock-outs

Health workers and managers in both districts described recurrent, sometimes unexplained stock-outs lasting several weeks. Caregivers returned home without supplies, CHWs faced hostility and limited stock was rationed. National and international participants described the upstream causes to be mostly related to procurement delays, bottlenecks in the national supply chain and constraints in last-mile distribution to health centres. Frontline respondents thus experienced the consequences of stock-outs that national participants explained, and both groups saw supply reliability as a main constraint on programme effectiveness.

## Discussion

This study compared how caregivers, health workers and health service managers experience community-based nutrition interventions in 2 contrasting settings in Rwanda. Three findings stand out. First, supplementation was universally valued, but its effect was diluted by household food insecurity and by families’ responses to eligibility and withdrawal rules. Second, CHWs held the system together while absorbing many of its failures, including hostility when commodities were unavailable. Third, the supply and financing constraints described by national participants explained much of what frontline respondents experienced.

### Supplementation in food-insecure households

Sharing of child-targeted supplements is well documented. In southern Ethiopia, caregivers treated therapeutic food both as medicine and as a household resource that could be shared or sold [33]. In Niger, researchers concluded that consumption by the target child alone is unrealistic where other household members are also hungry [34], and a recent Rwandan study reported the same practice for Shisha Kibondo [12]. This further explains the study participants experience in sharing food supplements mainly as a response to food insecurity.

Our findings add evidence of behavioural responses to the programme’s eligibility and exit rules, including early cessation of breastfeeding, avoidance of family planning and relapse after food support ended. In Ethiopia, some caregivers sought repeated admissions to prolong access to therapeutic food [33]. Because eligibility depends on the child’s age and nutritional status and on the household’s socio-economic category, a child-targeted benefit that becomes an important household resource creates incentives that can work against programme aims. Combining supplementation with household-level nutrition-sensitive support, such as social protection or livelihood interventions [35], and basing the end of food support on verified household circumstances, with a planned transition rather than an abrupt exit, could reduce these effects.

The competing explanations of stunting point in the same direction. Some frontline workers attributed persistent stunting to caregiver knowledge, whereas others, and national participants, emphasised land, livelihoods, flooding and time. Women’s workload is a recurring reason why agricultural and income gains fail to improve child nutrition [36], and women’s time constraints were the most frequently reported barriers to child health care as seen in northern Kenya [37]. Also, caregivers in South Africa linked malnutrition to poverty [25]. That income from cross-border trade or domestic work did not translate into caregiving time suggests that counselling alone is insufficient and that childcare arrangements deserve more attention in nutrition strategies.

### Community Health Workers as backbone and shock absorber

The centrality of CHWs, and the pressures on them, echo earlier Rwandan evidence of respected and motivated CHWs facing an overwhelming workload [38]; our findings suggest that this workload persists, together with shortages of basic anthropometric equipment. Remuneration, supervision and a reliable supply of commodities are key determinants of CHW performance [39,40], and the World Health Organization recommends that CHWs be remunerated in line with their workload [41]. Low pay and high turnover is common [40,42,43], however, were not prominent in our data, may reflect Rwanda’s cooperative and performance-based model [38] or CHWs’ reluctance to express dissatisfaction in discussions held near health facilities.

The safety concerns raised by CHWs in Bugeshi add a less-documented dimension, as regional research has mostly framed CHWs challenges in terms of workload and resources [26]. Because the delivery model links a CHW’s credibility to commodity availability, frustration falls on the CHWs when caregivers are mobilised to a facility that has nothing to give. Supply reliability is therefore a matter of workforce protection as well as service quality.

Accounts of CHWs declining referrals and under-reporting illustrate frontline discretion. Lipsky’s concept of street-level bureaucracy describes how frontline workers facing high demand and limited resources develop their own routines for rationing services [44], and such discretion has been shown to shape access to care in African health systems [45]. In Rwanda, performance targets may add a reputational motive: community members elsewhere perceived some programmes as implemented mainly to meet local leaders’ performance contract (“imihigo”) targets [12]. Discretion also allowed CHWs to deliver supplements privately to families who feared stigma, but it means that routine coverage indicators cannot show who was excluded at the frontline. Because declined referrals, perceived favouritism and under-reporting were reported by caregivers but are invisible in routine indicators, caregiver feedback is one of the few ways to detect them.

### Who is reached and who is missed

Eligibility for food supplementation rests on the child’s age and nutritional status and on the household’s socio-economic category. Through a horizontal equity lens, the gaps described were therefore mostly exclusion of children who met these criteria: rationing by distance during shortages, discretionary referral and enrolment, and poor households hidden in wealthier neighbourhoods [31,32]. Such exclusion bias is common in targeted programmes that rely on visible markers of poverty or on local administrators to identify beneficiaries [46], and misclassification and favouritism in beneficiary selection have been reported elsewhere in Rwanda [12]. Also, the exclusion of poor households in wealthier neighbourhoods in Rugarama is consistent with evidence that socio-economic inequalities in child stunting are larger within urban than within rural areas [47]. Therefore, screening of all children in a particular village, regardless of apparent wealth, can be a practical response.

Stigma may add a further barrier. In northern Kenya, caregivers of acutely malnourished children were considerably more likely than others to report shame as a barrier to care [37]. Fear of judgement also been described among caregivers of vulnerable infants in Malawi [24]. In this study, the participants described shame and concealment while denying that stigma existed fits a broader understanding of stigma as labelling and loss of status, not only overt discrimination [48]. Programmes that assess stigma only by asking about discrimination may therefore underestimate it.

Female caregivers gave more critical accounts of programme barriers, consistent with Rwandan evidence that household food allocation and spending decisions are gender biased [49]. Building on Rwanda’s gender-transformative programmes delivered by CHWs [50], community feedback on nutrition programmes should include separate spaces for women, so that the barriers they face are not underrepresented in mixed or male-dominated forums.

### Supply chains and financing

Stock-outs linked many of the problems described by frontline respondents. The reported coverage falls and recovering with stock availability suggests that coverage may partly reflect the availability of supplies rather than changes in children’s needs or in programme performance. Coverage assessments in 21 countries found that most community-based malnutrition treatment programmes did not reach recommended coverage, underlining the importance of measuring who is actually reached rather than relying on administrative figures [51]. However, local production of food supplements can shorten lead times and support local economies [52].

Dependence on time-limited donor funding was nuanced at community level through the loss of valued activities [12]. National participants proposed more predictable, multi-year and increasingly domestic financing. This need is becoming more pressing: official development assistance from members of the Organisation for Economic Co-operation and Development fell by an estimated 23% in real terms between 2024 and 2025, with the largest reduction in assistance from the United States [53]. Sustaining community-based nutrition services in this environment will depend on the extent to which domestic resources can replace declining external funding.

### Interpreting the contrast between the 2 sites

Alongside the analysis of routine screening data that informed site selection [8], the study findings suggest several hypotheses for the contrasting trajectories in Rubavu and Nyarugenge. In Bugeshi, participants described insecure access to land, water scarcity and related infection risks, also examined at district level in a companion ecological analysis [54], as well as flooding and time pressures linked to cross-border livelihoods. While in Rugarama, participants described a more urban and potentially better-resourced service environment, but also poor households remaining invisible in wealthier neighbourhoods. These factors are consistent with the differing trajectories but cannot, on qualitative evidence alone, be shown to have caused them, and should be tested in mixed methods implementation research.

### Implications for programme implementation

The study findings point to 7 practical steps: (1) confirming commodity availability before CHWs mobilise caregivers, to protect CHWs from the consequences of supply failure; (2) reviewing the rules for ending food support so that eligibility is based on verified household circumstances and is prepared in advance, and linking households whose children left the programme, or in which supplements are routinely shared, to provide social protection and livelihood support; (3) collecting structured, confidential caregiver feedback, including in separate spaces for women and men, to reveal problems that routine indicators miss, such as declined referrals, perceived favouritism and under-reporting; (4) equipping CHWs with adequate anthropometric equipment and protecting them from hostility when commodities are unavailable; (5) conducting targeted spot checks of routine data; (6) providing more predictable multi-year financing, with an increasing domestic share; and (7) assessing the feasibility and cost-effectiveness of decentralised production for each product.

### Study strength and limitations

A strength of this study is its comparison of accounts of the same interventions across 3 stakeholder groups, from caregivers to national programme managers, and in sites selected on trends quantified in an analysis of national surveillance data, so that the findings can be interpreted in relation to documented differences in outcomes. However, several limitations should be considered. Fieldwork was carried out in 1 health centre catchment area in each of the 2 districts, so the findings are not representative of all Rwanda. Although they may be transferable to similar settings; national interviews broadened the system perspective but do not substitute for additional sites. With 1 FGD per stakeholder group, sex and site, some issues may not have reached saturation within the FGD data, and comparisons between female and male caregivers are exploratory. Social desirability may have influenced discussions held in or near health facilities, although the substantial disagreement across groups and sites suggests that accounts were not uniformly positive.

## Conclusions

Community-based nutrition programmes in Bugeshi and Rugarama were broadly valued, and their rationale was well understood by caregivers, health workers and managers alike. Beneath this shared narrative, however, participants described an uneven implementation reality, supply disruptions felt most directly by CHWs, household food insecurity undermining the child-specific use of supplements, eligible children missed in both the rural and the urban setting, and frontline discretion that could both ease and obstruct access.

The findings from this study offer plausible, but not causal, explanations for the divergent stunting trajectories in Rubavu and Nyarugenge. Livelihoods, caregiving time, water and sanitation, urban inequality, supply reliability and frontline responsiveness may all shape how national policies translate into community-level outcomes, which are difficult to capture through routine surveillance alone.

Sustained progress will therefore require more than maintaining commodity coverage. It will require support for the CHWs who deliver nutrition services, attention to the household economic and caregiving conditions that shape their use, eligibility and withdrawal rules that reflect household circumstances, reliable supply and financing, and routine use of caregiver, health worker and manager perspectives to detect gaps between reported performance and lived experience.

## Data Availability

The datasets used and/or analyzed during the current study are available from the corresponding author on reasonable request.

## Acknowledgments

The authors thank the participants for their time and candour and note their explicit request that study findings be shared with their communities and district authorities. The authors also thank the CMHS Institutional Review Board, University of Rwanda, for its review of the study.

## Supporting information

**S1 Checklist. Completed COREQ checklist.**

**S2 Checklist. Inclusivity in global research questionnaire.**

**S3 Semi-structured topic guides for caregiver FGDs, CHW FGDs and in-depth interviews.**

## Notes

### Competing Interest Statement

The authors have declared no competing interest.

### Author Declarations

The study was approved by the Institutional Review Board of the College of Medicine and Health Sciences, University of Rwanda (Approval Notice No. 452/CMHS IRB/2026 approved 27 April 2026). All participants gave written informed consent, and transcripts were de-identified before analysis.

## References

1. Black RE, Victora CG, Walker SP, Bhutta ZA, Christian P, De Onis M, et al. Maternal and child undernutrition and overweight in low-income and middle-income countries. The lancet. 2013;382: 427–451.

2. Economic Development and Poverty Reduction Strategy II. Available: https://www.kigalicity.gov.rw/fileadmin/user_upload/Kigali_city/Publications/Policies/Economic_Development_and_Poverty_Reduction_Strategy2__2013-2018.pdf

3. 7 Years Government Programme: National Strategy for Transformation (NST1) 2017-2024. Available: https://faolex.fao.org/docs/pdf/rwa206814.pdf

4. Republic of Rwanda, Ministry of Finance and Economic Planning. Vision 2050. Available: https://www.minecofin.gov.rw/fileadmin/user_upload/Minecofin/Publications/REPORTS/National_Development_Planning_and_Research/Vision_2050/English-Vision_2050_Abridged_version_WEB_Final.pdf

5. Five Years Government Programme: Rwanda National Strategy for Transformation (NST2) 2024-2029. Available: https://www.minecofin.gov.rw/index.php?eID=dumpFile&t=f&f=112650&token=cb55b3319372c3f73528c46433b587ef72e8d4eb

6. National Institute of Statistics Rwanda. Rwanda Demographic and Health Survey 2019-20. 2020. Available: https://www.statistics.gov.rw/publication/1994

7. National Institute of Statistics of Rwanda. Rwanda Demographic and Health Survey (2025). 2025. Available: https://statistics.gov.rw/data-sources/surveys/Demographic-and-Health-Survey/demographic-and-health-survey-2025/demographic-and-health-survey-2025-final-report

8. Ntawuyirushintege S, Ahmed A, Bucyibaruta G, Siddig EE, Remera E, Tediosi F, et al. Spatiotemporal Trends in Stunting Prevalence Among Children Aged Two Years Old in Rwanda (2020–2024): A Retrospective Analysis. Nutrients. 2025;17: 2808.

9. Republic of Rwanda National Food and Nutrition Policy 2013-2018. Available: https://www.moh.gov.rw/fileadmin/user_upload/policies/National_Food_and_Nutrition_Policy_.pdf

10. Republic of Rwanda National Food and Nutrition Strategic Plan 2013-2018. Available: https://www.policyvault.africa/wp-content/uploads/policy/RWA310.pdf

11. Republic of Rwanda Fouth Health Sector Strategic Plan July 2018-June 2024. Available: https://www.moh.gov.rw/fileadmin/user_upload/Moh/New_publication/Srategic-plans-priorities/Fouth_Health_Sector_Strategic_Plan__HSSP4__July_2018-June_2024.pdf

12. Mporanyi T, Absolomon G, Rubuga FK, Condo J, Munyanshongore C. Assessing community awareness, perceptions, ownership, and challenges of stunting reduction programs in Rwanda: a qualitative study. BMC Public Health. 2026;26: 1496.

13. Mogojwe H. Rwanda’s groundbreaking food supplement venture slashes child malnutrition, sets global benchmark. In: Clinton Health Access Initiative [Internet]. 29 May 2024 [cited 19 Oct 2026]. Available: https://www.clintonhealthaccess.org/blog/chronic-malnutrition-declines-in-rwanda-thanks-to-fortified-blended-foods/

14. THE ASSESSMENT OF THE IMPACT OF HOME GROWN INITIATIVES. Available: https://www.rgb.rw/fileadmin/user_upload/RGB/Publications/HOME_GROWN_SOLUTIONS/Impact_of_Home_Grown_Initiative_2014.pdf

15. Citizens start classifying their households into new Ubudehe categories. [cited 23 Oct 2026]. Available: https://www.loda.gov.rw/news-detail/citizens-start-classifying-their-households-into-new-ubudehe-categories-1

16. Nkurunziza M. What next after govt scraps Ubudehe categories? In: The New Times [Internet]. 31 Mar 2023 [cited 23 Oct 2026]. Available: https://www.newtimes.co.rw/article/6292/news/rwanda/what-next-after-govt-scraps-ubudehe-categories

17. Cabinet approves review and classification of households into Ubudehe categories. [cited 19 Oct 2026]. Available: https://www.loda.gov.rw/news-detail/cabinet-approves-review-and-classification-of-households-into-ubudehe-categories-1

18. Imibereho Dynamic Social Registry launched. 8 Oct 2026 [cited 23 Oct 2026]. Available: https://www.minaloc.gov.rw/news-detail/imibereho-dynamic-social-registry-launched

19. CBHI / Mutuelle de Sante | RSSB Rwanda. [cited 23 Oct 2026]. Available: https://rssb.rw/schemes/cbhi-scheme

20. Uwera T, Frøen JF, Papadopoulou E, Rukundo E, Sibomana H, Muhire A, et al. Child immunization data quality in Rwanda: an assessment of routine health information system data. Archives of Public Health. 2025;83: 97.

21. GIKURIRO Program - The Integrated Nutrition and WASH Activity Baseline Survey Report March 2017. Available: https://rbc.gov.rw/compass/wp-content/uploads/gikuriro_program_baseline_survey.pdf

22. World Bank. Rwanda - Stunting Prevention and Reduction Project: environmental assessment: Medical waste management plan. In: World Bank [Internet]. [cited 19 Oct 2026]. Available: https://documents.worldbank.org/en/publication/documents-reports/documentdetail/659321512155398462

23. World Bank. Development Projects: Additional Financing to Rwanda Stunting Prevention and Reduction Project - P179499. In: World Bank [Internet]. [cited 19 Oct 2026]. Available: https://projects.worldbank.org/en/projects-operations/project-detail/P179499

24. Koenraads M, Phuka J, Maleta K, Theobald S, Gladstone M. Understanding the challenges to caring for low birthweight babies in rural southern Malawi: a qualitative study exploring caregiver and health worker perceptions and experiences. BMJ global health. 2017;2: e000301.

25. Thabathi TE, Maluleke M, Raliphaswa NS, Masutha TC. Perceptions of caregivers regarding malnutrition in children under five in rural areas, South Africa. Children. 2022;9: 1784.

26. Wambui E, Wilunda C, Donfouet HPP, Mwangi B, Zerfu TA, Daniel T, et al. Perceptions towards management of acute malnutrition by community health volunteers in northern Kenya. PLOS global public health. 2024;4: e0002564.

27. Braun V, Clarke V. Using thematic analysis in psychology. Qualitative research in psychology. 2006;3: 77–101.

28. Gale NK, Heath G, Cameron E, Rashid S, Redwood S. Using the framework method for the analysis of qualitative data in multi-disciplinary health research. BMC medical research methodology. 2013;13: 117.

29. Tong A, Sainsbury P, Craig J. Consolidated criteria for reporting qualitative research (COREQ): a 32-item checklist for interviews and focus groups. International journal for quality in health care. 2007;19: 349–357.

30. Guest G, Bunce A, Johnson L. How many interviews are enough? An experiment with data saturation and variability. Field methods. 2006;18: 59–82.

31. Whitehead M. The concepts and principles of equity and health. International journal of health services. 1992;22: 429–445.

32. Culyer AJ, Wagstaff A. Equity and equality in health and health care. Journal of health economics. 1993;12: 431–457.

33. Tadesse E, Berhane Y, Hjern A, Olsson P, Ekström E-C. Perceptions of usage and unintended consequences of provision of ready-to-use therapeutic food for management of severe acute child malnutrition. A qualitative study in Southern Ethiopia. Health Policy and Planning. 2015;30: 1334–1341.

34. Marquer C, Langendorf C, Woi-Messe LC, Berthe F, Ategbo E-A, Rodas-Moya S, et al. Intrahousehold management and use of nutritional supplements during the hunger gap in Maradi region, Niger: a qualitative study. BMC nutrition. 2020;6: 4.

35. Ruel MT, Alderman H. Maternal and Child Nutrition Study Group Nutrition-sensitive interventions and programmes: how can they help to accelerate progress in improving maternal and child nutrition. Lancet. 2013;382: 536–51.

36. Johnston D, Stevano S, Malapit HJ, Hull E, Kadiyala S. Time use as an explanation for the agri-nutrition disconnect: evidence from rural areas in low and middle-income countries. Food policy. 2018;76: 8–18.

37. Bliss JR, Njenga M, Stoltzfus RJ, Pelletier DL. Stigma as a barrier to treatment for child acute malnutrition in M arsabit C ounty, K enya. Maternal & child nutrition. 2016;12: 125–138.

38. Condo J, Mugeni C, Naughton B, Hall K, Tuazon MA, Omwega A, et al. Rwanda’s evolving community health worker system: a qualitative assessment of client and provider perspectives. Human resources for health. 2014;12: 71.

39. Kok MC, Dieleman M, Taegtmeyer M, Broerse JE, Kane SS, Ormel H, et al. Which intervention design factors influence performance of community health workers in low-and middle-income countries? A systematic review. Health policy and planning. 2015;30: 1207–1227.

40. Scott K, Beckham SW, Gross M, Pariyo G, Rao KD, Cometto G, et al. What do we know about community-based health worker programs? A systematic review of existing reviews on community health workers. Human resources for health. 2018;16: 39.

41. World Health Organization. WHO guideline on health policy and system support to optimize community health worker programmes. World Health Organization; 2018.

42. Ngabonziza JCS, Suleiman K, Mohamed NS, Rukundo G, Muremba MF, Kagame J, et al. Implementation and evaluation of a community health workers-led digital integrated diseases screening system to provide healthcare for patients at community level in Rwanda. EBioMedicine. 2026;125: 106168. doi:10.1016/j.ebiom.2026.106168

43. Ngabonziza JCS, Mohamed NS, Muremba MF, Rutebuka H, Mugumya S, Gihana MW, et al. Digitally Integrated Disease Management System in Rwanda: Community Healthcare Worker Perspectives. The American journal of tropical medicine and hygiene. 2026; tpmd260053.

44. Lipsky M. Street-level bureaucracy: Dilemmas of the individual in public service. Russell sage foundation; 2010.

45. Walker L, Gilson L. ‘We are bitter but we are satisfied’: nurses as street-level bureaucrats in South Africa. Social science & medicine. 2004;59: 1251–1261.

46. Coady D, Grosh ME, Hoddinott J. Targeting of transfers in developing countries: Review of lessons and experience. 2004.

47. Menon P, Ruel MT, Morris SS. Socio-economic differentials in child stunting are consistently larger in urban than in rural areas. Food and Nutrition Bulletin. 2000;21: 282–289.

48. Link BG, Phelan JC. Conceptualizing stigma. Annual review of Sociology. 2001;27: 363–385.

49. Albin MQ, Igihozo G, Musemangezhi S, Namukanga EN, Uwizeyimana T, Alemayehu G, et al. “When we have served meat, my husband comes first”: A qualitative analysis of child nutrition among urban and rural communities of Rwanda. Plos one. 2024;19: e0306444.

50. Doyle K, Bhatnagar I, Karamage E, Tuyisingize JP, Muhimpundu C, Nyiransabimana AMY, et al. Equipping community health workers in Rwanda to deliver a gender transformative parenting program to prevent violence against women and children at scale. Frontiers in reproductive health. 2025;7: 1602136.

51. Rogers E, Myatt M, Woodhead S, Guerrero S, Alvarez JL. Coverage of community-based management of severe acute malnutrition programmes in twenty-one countries, 2012-2013. PloS one. 2015;10: e0128666.

52. Segrè J, Liu G, Komrska J. Local versus offshore production of ready-to-use therapeutic foods and small quantity lipid-based nutrient supplements. Maternal & Child Nutrition. 2017;13: e12376.

53. International aid fell sharply in 2025, says OECD. [cited 19 Oct 2026]. Available: https://www.oecd.org/en/about/news/press-releases/2026/04/international-aid-fell-sharply-in-2025-says-oecd.html

54. Ntawuyirushintege S, Mohamed NS, Ahmed A, Semakula M, Remera E, Tediosi F, et al. District-level ecological analysis of the association between stunting and infectious diseases among children under 2 years in Rwanda, 2020–2024. Frontiers in Public Health. 2026;14: 1855873.

